# Traditional surveys versus ecological momentary assessments: digital citizen science approaches to improve ethical physical activity surveillance among youth

**DOI:** 10.1101/2023.06.06.23291067

**Authors:** Sheriff Tolulope Ibrahim, Nour Hammami, Tarun Reddy Katapally

## Abstract

**Background:** The role of physical activity (PA) in minimizing non-communicable diseases is well established. Measurement bias can be reduced via ecological momentary assessments (EMAs) deployed via citizen-owned smartphones. This study aims to engage citizen scientists to understand how PA reported digitally by retrospective and prospective measures varies within the same cohort.

**Methods:** This study used the digital citizen science approach to collaborate with citizen scientists, aged 13-21 years over eight consecutive days via a custom-built app. Citizen scientists were recruited through schools in Regina, Saskatchewan, Canada in 2018 (August 31 - December 31). Retrospective PA was assessed through a survey, which was adapted from three validated PA surveys to suit smartphone-based data collection, and prospective PA was assessed through time-triggered EMAs deployed consecutively every day, from day 1 to day 8, including weekdays and weekends. Data analyses included t-test to understand the difference in PA reported retrospectively and prospectively, and linear regressions to assess contextual and demographic factors associated with PA reported retrospectively and prospectively.

**Result:** Findings showed a significant difference between PA reported retrospectively and prospectively (p = 0.001). Ethnicity (visible minorities: β = - 0.911, 95% C.I.= -1.677, -0.146), parental education (university: β = 0.978, 95% C.I.= 0.308, 1.649), and strength training (at least one day: β = 0.932, 95% C.I.= 0.108, 1.755) were associated with PA reported prospectively. In contrast, the number of active friends (at least one friend: β = 0.741, 95% C.I.= 0.026, 1.458) was associated with retrospective PA.

**Conclusion:** Physical inactivity is the fourth leading cause of mortality globally, which requires accurate monitoring to inform population health interventions. In this digital age, where ubiquitous devices provide real-time engagement capabilities, digital citizen science can transform how we measure behaviours using citizen-owned ubiquitous digital tools to support prevention and treatment of non-communicable diseases.

**Author summary:** Traditionally, the surveillance of physical activity has been predominantly conducted with retrospective surveys that require participants to recall behaviours, a methodology which has significant challenges due to measurement bias. With advances in digital technology, ubiquitous devices offer a solution through ecological momentary assessments (EMAs). Using the Smart Framework, which combines citizen science with community-based participatory research, this study ethically obtained retrospective and prospective EMA physical activity data from the same cohort of youth citizen scientists, who used their own smartphones to engage with our team over an eight-day period. The findings show a significant difference between physical activity reported through retrospective and prospective EMAs. Moreover, there was also a variation between contextual and demographic factors that were associated with retrospective and prospective physical activity – evidence that points towards the need to adapt physical activity surveillance in the digital age by ethically engaging with citizens via their own ubiquitous digital devices.

## Background

Physical activity is an important protective factor that can prevent or minimize non-communicable diseases such as diabetes mellitus, cancer, obesity, hypertension, and joint conditions [1–4]. However, measuring physical activity (PA) can be plagued with challenges and inaccuracies, such as over-reporting of PA [5], recall bias [6], lack of environmental and social context of PA, and difficulty in reporting PA in the form of intensities (e.g., moderate, and vigorous activities) [7–9]. The continued understanding of how PA is accumulated, and its accurate measurement, is crucial in identifying patterns of PA, and to accurately monitor population adherence to PA recommendations [10,11].

Traditionally, retrospective means of measurement have been used to measure PA accumulation [12,13]. Retrospective surveys in general are easy to implement and are not resource intensive [14], however they tend to overestimate PA [15,16], which can be attributed to recall biases [8,17]. For instance, in a study carried out on lower back pain patients and healthy controls, it was determined that retrospective surveys overestimated self-reported moderate physical activity by 42min/day, and vigorous activity by 39min/day [18]. Objective measures of PA, such as the use of accelerometers, global positioning system, heart rate monitoring, and movement sensors, can solve the problem of recall bias present in retrospective subjective questionnaires. However, objective measures can be time consuming [19], expensive [20], and logistically challenging to implement across populations [21–23].

Advances in information and communication technology offer novel opportunities in PA measurement [24,25]. Ubiquitous tools such as smartphones can enable ecological momentary assessments (EMAs) to be deployed via smartphones in near real-time, and with more frequency by using time-, location-, and user-triggers, which provide flexibility for both researchers and study participants [26,27]. EMAs assess participants’ experience/behaviour in real-time, and in the real-world, where researchers use sampling and monitoring strategies to assess phenomena as they occur in natural settings [17]. More recently, the use of EMAs in assessing PA behaviour using citizen-owned smartphones has gained momentum among researchers [26–28], due to its ability to eliminate recall [23], and social desirability biases [29] that are inherent in retrospective subjective PA surveys [30,31].

This study aims to ascertain if there is a significant difference between the duration of PA reported retrospectively using traditional validated surveys, and duration of PA reported prospectively using EMAs, within the same cohort of participants. In addition, this study assesses contextual and demographic factors that are associated with duration of PA reported retrospectively vs. prospectively (EMAs) within the same cohort of participants.

## Methodology

This study captures PA behaviours, and its related factors from youth who participated in the Smart Platform as youth citizen scientists [26]. The Smart Platform is a citizen science and digital epidemiological initiative for population health surveillance, knowledge translation, and real-time interventions [32,33]. It combines participatory, community-based, and citizen science approaches to leverage citizen-owned smartphones to ethically engage citizen scientists for population health research. The research ethics approval for the Smart Platform was approved by the Research Ethics Boards of the Universities of Regina and Saskatchewan (REB # 2017-029).

The Smart Platform enabled our research team to use a custom-built smartphone application (app) to engage with citizen scientists [26,27] over eight consecutive days [34] Youth citizen scientists had the option to download the app from both the iOS and Android platforms onto their smartphones. Using the app, apart from PA data, a wide range of behavioural, contextual, demographic, social factors were reported by youth [26,27,33]. This study used the following data that were derived using surveys deployed via the app [25,26]: family and peer support for PA; sociodemographic characteristics; and individual characteristics that determined overall PA, such as strength training.

## Participants

A total of 808 youth citizen scientists (13 – 21 years) were recruited for this study. Youth citizen scientists were recruited through Regina Public and Catholic Schools engagement sessions held in various high schools in Regina, Saskatchewan, Canada in 2018. Citizen scientists were recruited through a collaborative effort between the school administrators and the research team. Scheduled in-person recruitment sessions were organized between the research team and the youth. Activities during the recruitment sessions included describing the study to youth, demonstration of how to use the app, answering queries and concerns, and assisting youth in downloading the app onto their respective smartphones. All youth participants of the study provided informed consent through the app. For youth participants between the ages of 13-16 years, implied informed consent was obtained from their caregivers and parents before the scheduled recruitment sessions.

## Measures

### PA (dependent variables)

On day one of the study, using a time-triggered smartphone nudge, retrospective PA data (over previous 7 days) were collected from youth through a modified survey adapted from three validated self-reported measures [35–39]. The modification allowed time-triggered digital deployment of the retrospective survey and accommodated the varying start dates of youth joining the study (**Figure 1**). Youth could download and join the study between (August 31 - December 31). Irrespective of the time when youth joined the study, a time-triggered retrospective survey was deployed that ensured the exact dates of the previous 7 days were reflected in the app to improve recall (**Figure 1**).

**Figure 1:**
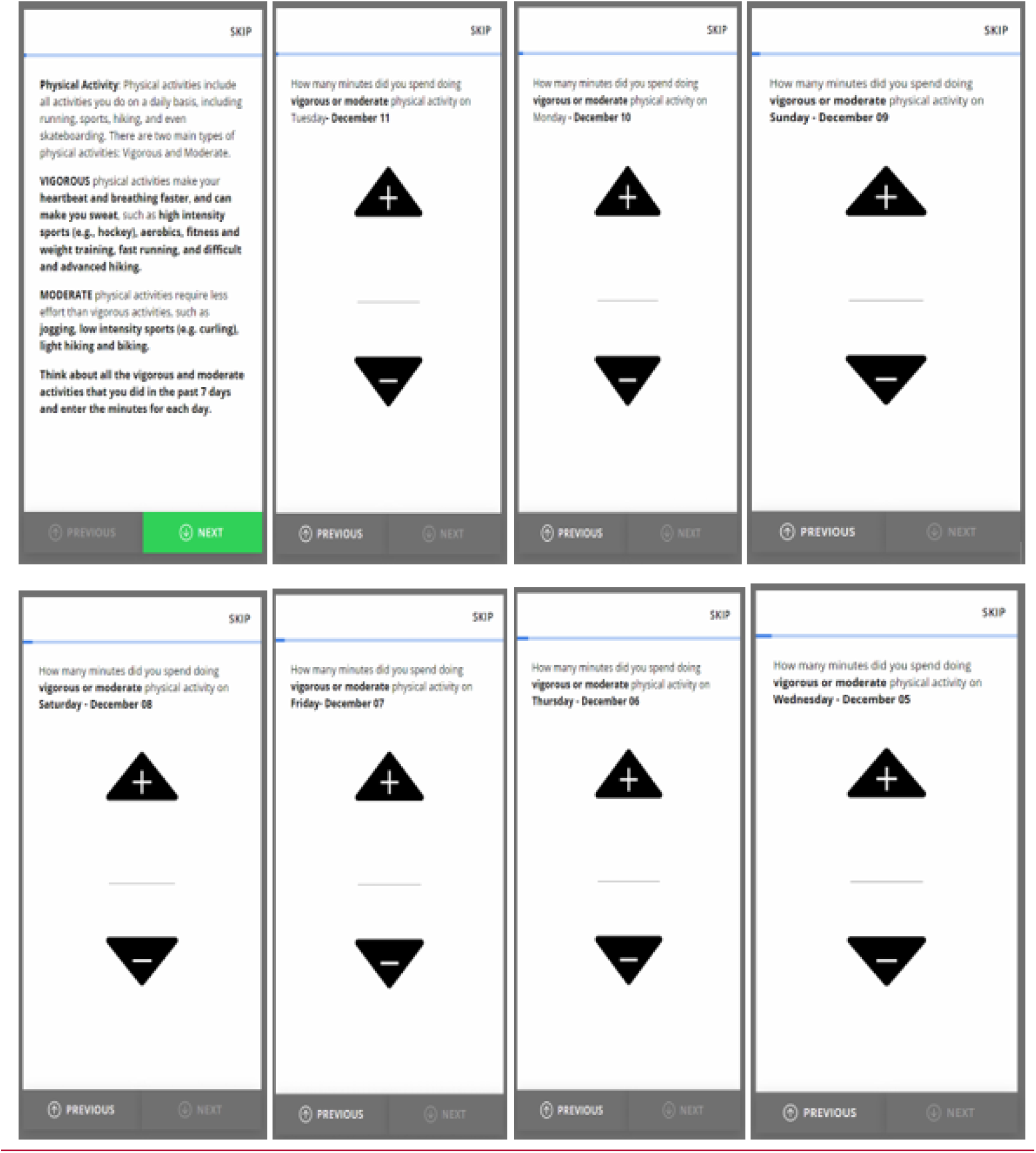
Digitally deployed modified retrospective PA survey.

After defining PA, the retrospective survey provided youth the opportunity to report PA they accumulated over the previous seven days starting from the day of joining the study. From these responses, mean overall PA per day (will be referred to as: retrospective PA duration) was derived. Following general PA data derivation standards [40,41], youth who reported less than 10 minutes or more than 960 minutes (about 16 hours) per day were excluded from the analyses [41,42].

Prospective PA information was obtained via daily time-triggered EMAs throughout the study period (eight consecutive days), to include both weekdays and weekends. The EMAs were deployed every evening between 8:00 PM to 11: 30 PM and were set to expire at midnight [26]. EMAs used skip-pattern questions to capture PA accumulation (**Figure 2**). After defining what constitutes PA, EMAs asked youth the following questions: 1) “What type of physical activities did you do today?” (Multiple choice); 2) “How many minutes did you spend doing this activity?” (Open ended). From these questions, mean PA per day was derived (will be referred to as: EMA PA duration).

**Figure 2:**
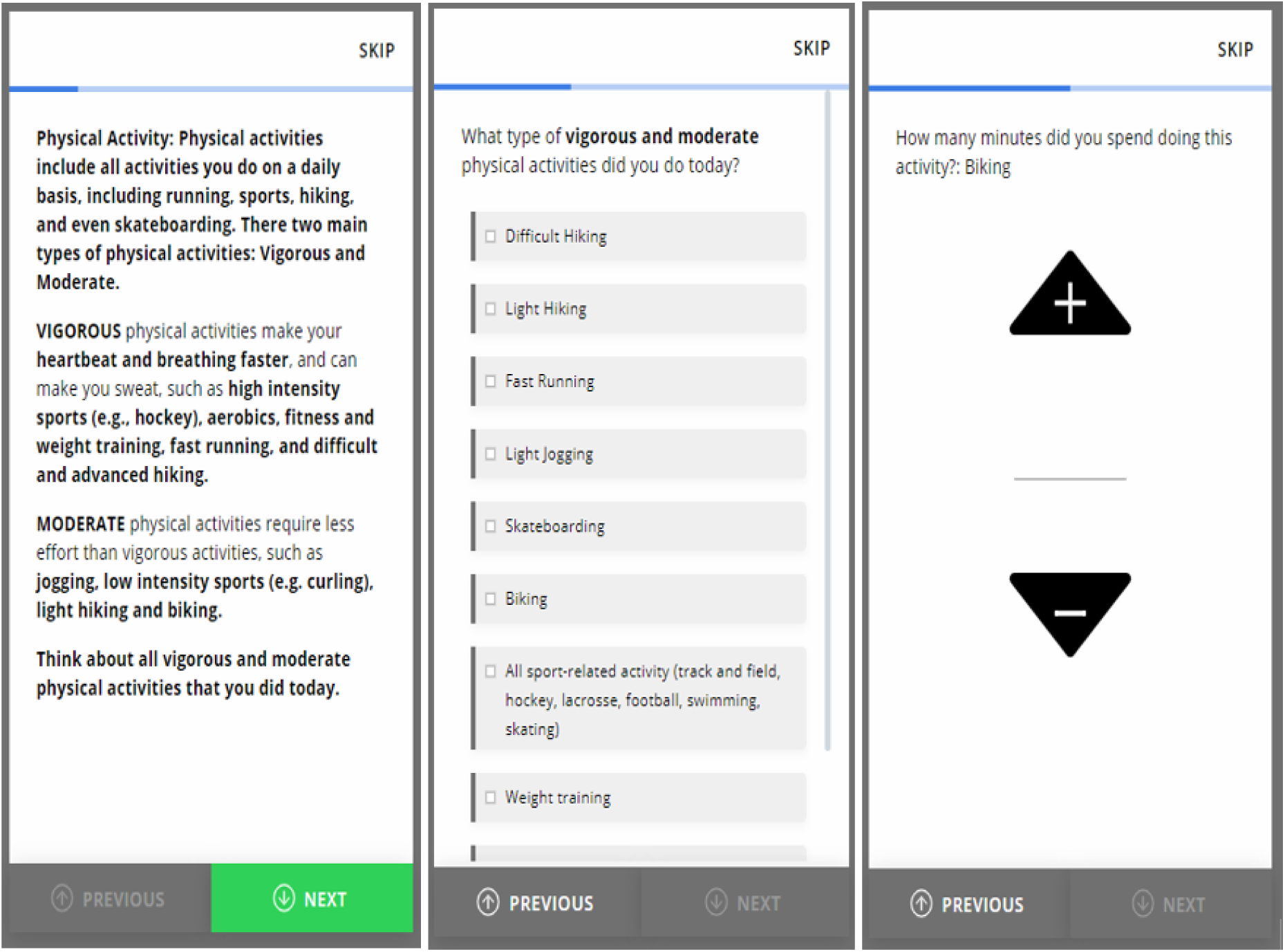
EMAs PA types, and duration.

### Family support for PA (independent variables)

Family support PA was captured using one question: “How much do your parents, stepparents, or guardians support you in being physically active? (e.g., driving you to team games, buying you sporting equipment)” with the 4 response options: “very supportive”, “supportive”, “unsupportive”, or “very unsupportive”. Using these data, we collapsed the responses into: “unsupportive” (combining unsupportive and very unsupportive), “supportive” and “very supportive”.

### Peer support for PA (independent variables)

Youth were asked to think about their closest friends in the last 12 months when answering the question regarding peer support of PA. Peer support for PA was captured with the question: “How many of your closest friends are physically active?” with the six response options: “none of my friends”, “1”, “2”, “3”, “4”, or “5 of my friends”. This variable was dichotomized into “0 physically active friends” corresponding to “none of my friends” and “at least 1 active friend” corresponding to “1”, “2”, “3”, “4” or “5 of my friends”.

### Sociodemographic covariates

Gender was ascertained with the question, “What is your gender?”, with 5 response options: “male”, “female”, “transgender”, “other (please specify)”, and “prefer not to disclose”. The responses “transgender”, “other”, and “prefer not to disclose” were collapsed into one category due to low counts within these categories. Parental education was measured by asking youth to report the “highest education” of one of their parents or guardians, with six response options: “elementary school”, “some secondary/high school”, “completed high school”, “some post-secondary (university/college)”, “received university or college degree /diploma”, and “does not apply”. From these responses, four categories of parental education were derived: 1) “elementary school” corresponds to “elementary school or below”, 2) “some secondary/high school” and “completed high school” corresponds to “at least secondary school” 3) “some post-secondary (university /college)”, “received university or college degree/diploma” corresponds to “university and above” and “does not apply”.

Youth citizen scientists were also asked about their ethnicity, with the following response options: “First Nations”, “Dene”, “Cree”, “Metis”, “Inuit”, “African”, “Asian”, “Canadian”, “Caribbean/West Indian”, “Eastern European”, “European”, “South Asian”, “other”, and “Mixed”. From these responses, four categories were extracted: 1) “Indigenous” which corresponds to “First Nations”, “Dene”, “Cree”, “Metis”, “Inuit”, 2) “Canadian”, 3) “mixed” and 4) “visible minorities”. The visible minorities include “African”, “Asian”, “Caribbean/West Indian”, “Eastern European”, “European”, “South Asian”, and “other” categories. The visible minorities category was created due to low count within these ethnic categories.

### Strength training (independent variables)

Strength training was captured using the following questions “On how many days in the last 7 days did you do exercises to strengthen or tone your muscles? (e.g., push-ups, sit-ups, or weight-training)” with the eight response options including “0”, “1”, “2”, “3”, “4”, “5”, “6”, or “7 days”. We dichotomized these responses into “0 days of strength training” corresponding to 0 days and “at least 1 day of strength training” corresponding to “0”, “1”, “2”, “3”, “4”, “5”, “6”, or “7 days”.

### Data and risk management

To ensure confidentiality, all data were encrypted before being streamed to a secure cloud server. Identifiable artifacts (e.g., photos, voice recordings) were removed or de-identified before the data were analyzed. A permission built into the app restricted access to personally identifiable information (e.g., contact list or network visited). Media Access Control address anonymization was used to protect youth citizen scientists’ data based on a simple hash algorithm. Risks and privacy management options were made clear to youth citizen scientists while obtaining informed consent. In addition, citizen scientists not only had the option to drop out of the study or pause data gathering anytime they wished, but also had the option to upload data only when they had Wi-Fi access and /or when their smartphones were plugged into a power source. Together with the above features, youth citizen scientists also had the option to drop out of the study at any point of time [27] **(Figure 3)**.

**Figure 3:**
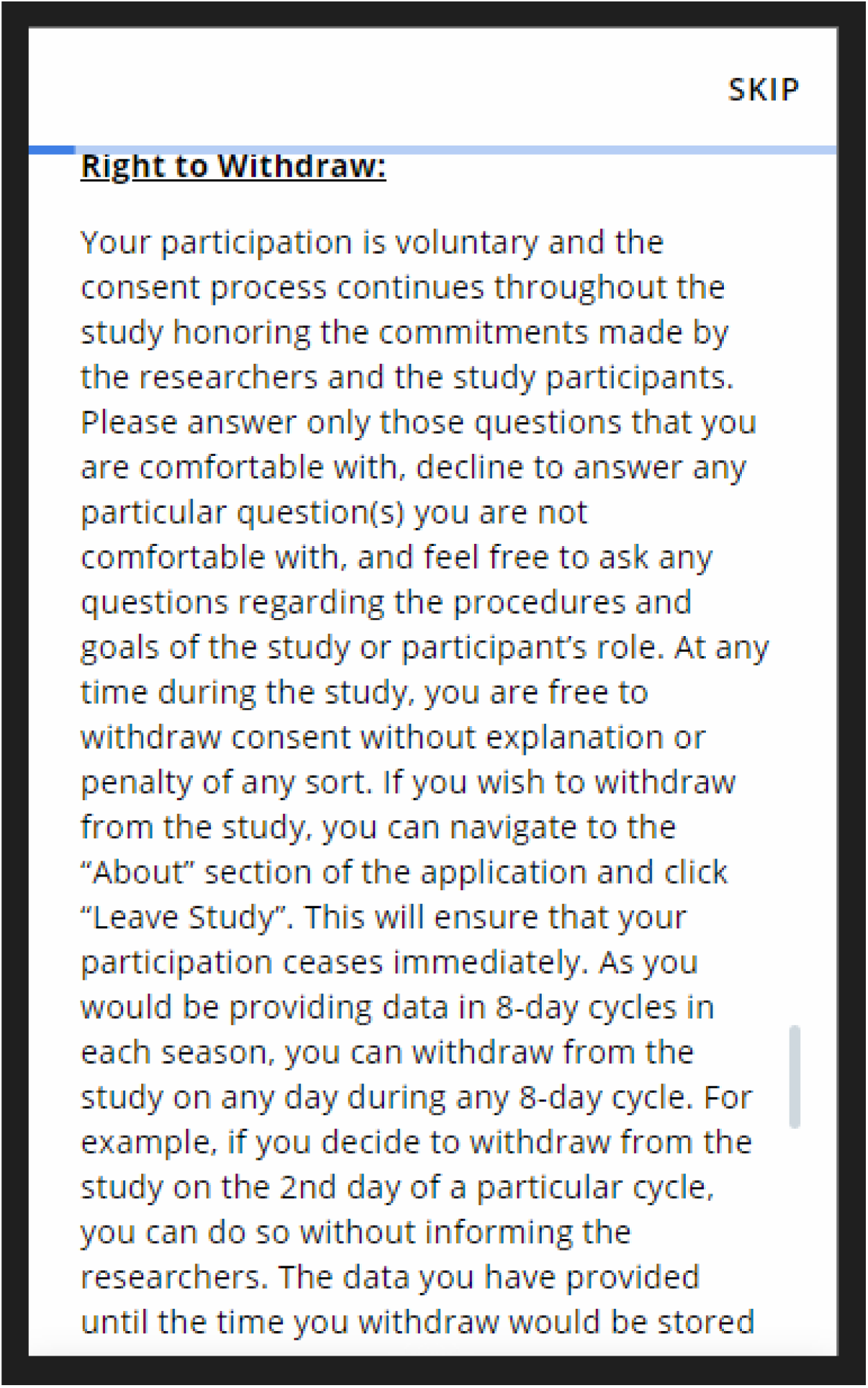
Study withdrawal information feature

## Data Analysis

Descriptive statistics, such as frequencies and percentages were used in describing the independent variables of this study. T-test inferential statistics was used to ascertain difference between mean minutes of PA reported via retrospective PA survey and mean minutes of PA reported via EMAs. Multiple linear regression models were used to assess factors associated with mean minutes of PA reported retrospectively and prospectively (EMAs), while adjusting for control variables. Data analyses were conducted using R 4.2.1 statistical tool. A significance level was set at p < 0.05.

## Results

A total of 808 youth citizen scientists (13 – 21 years) were recruited for this study. After excluding participants who did not report on primary dependent and independent variables, the final sample size of this study was 436.

Table 1 presents the summary statistics of youth citizen scientists. Youth were predominantly females (55.8%), with 38.5% being males and 5.7% reporting one of following categories: transgender, other, or preferred not to disclose. Majority of youth identified themselves as Canadian (39.8%), followed by mixed (29.7%), visible minority (25.5%), and Indigenous (5%). In terms of socioeconomic status, most youth (65.1%) reported that one of their parents had a university degree. In terms of strength training, 76.5% of youth reported having at least one day of strength training, while 23.5% reported having zero days of strength training.

**Table 1:** Summary statistics for youth citizen scientists participating in this study (n=436).

|  | Percentage |
| --- | --- |
| **Gender** |  |
| Male (n=161) | 38.5 |
| Female (n=233) | 55.8 |
| Transgender / Other / Prefer not to disclose (n=24) | 5.7 |
| Total (n=418) <sup>a</sup> | 100 |
| **Ethnicity** |  |
| Indigenous (n=21) | 5.0 |
| Canadian (n=166) | 39.8 |
| Mixed (n= 124) | 29.7 |
| Visible minority (n=106) | 25.5 |
| Total (n=417) <sup>a</sup> | 100 |
| **School** |  |
| 1 (n=110) | 25.3 |
| 2 (n=74) | 17.1 |
| 3 (n=50) | 11.5 |
| 4 (n=78) | 18.0 |
| 5 (n=122) | 28.1 |
| Total (n=434) <sup>a</sup> | 100 |
| **Grade** |  |
| Grade 9 (n=125) | 29.7 |
| Grade 10 (n=86) | 20.4 |
| Grade 11 (n=61) | 14.5 |
| Grade 12 (n=149) | 35.4 |
| Total (n=421) <sup>a</sup> | 100 |
| **Parental education** |  |
| Elementary or below (n= 12) | 2.8 |
| At least secondary school (n=91) | 21.0 |
| University and above (n =282) | 65.1 |
| Does not apply (n=48) | 11.1 |
| Total (n=433) <sup>a</sup> | 100 |
| **Strength training** |  |
| zero days of strength training (n=97) | 23.5 |
| At least one day of strength training (n=315) | 76.5 |
| Total (n=412) <sup>a</sup> | 100 |
| **Family support for PA** |  |
| Unsupportive (n=49) | 11.8 |
| Supportive (n=188) | 45.3 |
| Very supportive (n=178) | 42.9 |
| Total (n=415) <sup>a</sup> | 100 |
| **Peer Support for PA** |  |
| zero active friends (n= 47) | 11.3 |
| At least one active friend (n=368) | 88.7 |
| Total (n=415)<sup>a</sup> | 100 |
| <sup>a</sup> Some youth did not provide response to this question |  |

In terms of social context/support for PA, 88.7% reported having at least one or more physically active friends, while 45.3% reported that parents/guardians are supportive of their PA, and 42.9% reported that parents/guardians are very supportive of their PA.

The summary statistics of the dependent variables: retrospective PA and prospective PA EMAs were presented in Table 2. The mean time spent on PA per day (in minutes) reported via the retrospective PA survey and prospective EMAs were 93 and 196, respectively. The t-test result of 3.237 (p = 0.001) suggests a statistically significant difference between mean duration of PA reported by youth retrospectively and prospectively (EMAs).

**Table 2:** Mean time spent on PA per day (in minutes) as reported in the retrospective survey and the prospective EMAs and t-test analysis.

|  | Mean<br>(Minutes per<br>day) | Minimum<br>(minutes) | Maximum<br>(minutes) | t-test (p-value) |
| --- | --- | --- | --- | --- |
| Retrospective PA<br>(Measured via<br>retrospective survey) | 93 | 13.0 | 557.0 | 3.237<br>(0.001) |
| Prospective PA<br>(Measured via EMA) | 196 | 10.0 | 910.0 |  |

The adjusted, linear regression models showing the relationship between (EMA PA [model 1] and Retrospective PA [model 2]), and contextual and demographic factors are presented in Table 3.

**Table 3:** Factors associated with PA duration measured via prospective (EMA; results of Model 1) and retrospective PA survey measures (Retrospective PA; results of Model 2).

|  | **Model 1: EMA PA**<br>**duration** |  | **Model 2: Retrospective**<br>**PA duration** |  |
| --- | --- | --- | --- | --- |
|  | **Beta**<br>**coefficients<sup>a</sup>**<br>**(95% CI)** | **P-value** | **Beta**<br>**coefficients<sup>a</sup>**<br>**(95% CI)** | **P-value** |
| **Ethnicity -Canadian (Ref.)** |  |  |  |  |
| Indigenous | -3.336 (-6.816, 0.143) | 0.066 | 0.163 (-2.726, 3.052) | 0.912 |
| Mixed | 0.287 (-0.350, 0.925) | 0.381 | 0.205 (-0.324, 0.734) | 0.451 |
| Visible minority | -0.911*** (-1.677, -0.146) | 0.024 | 0.106 (-0.530, 0.742) | 0.745 |
| **Parental education<sup>b</sup> – At least secondary school (Ref.)** |  |  |  |  |
| University and above | 0.978*** (0.308, 1.649) | 0.006 | 0.262 (-0.294, 0.819) | 0.360 |
| Does not apply | 0.768 (-0.341, 1.878) |  | -0.016 (-0.937, -0.905) | 0.973 |
| **Strength training** |  |  |  |  |
| **zero days of strength training (Ref.)** |  |  |  |  |
| At least one day of strength training | 0.932*** (0.108, 1.755) | 0.031 | 0.357 (-0.326, 1.041) | 0.310 |
| **Family support for PA** |  |  |  |  |
| **Unsupportive (Ref.)** |  |  |  |  |
| Supportive | -0.593 (-1.479, -0.293) | 0.195 | -0.702 (-1.438, 0.033) | 0.067 |
| Very supportive | -0.412 (-1.253, 0.429) | 0.342 | -0.026 (-0.724, 0.673) | 0.942 |
| **Peer support for PA** |  |  |  |  |
| **zero physically active friends (Ref.)** |  |  |  |  |
| At least one active friend | 0.169 (-1.693, 1.032) | 0.702 | 0.741*** (0.026, 1.458) | 0.048 |
| **Constant** | 4.215 (-1.496,<br>9.927) | 0.154 | 1.339 (-3.402,<br>6.081) | 0.582 |
| *** $p < 0.05$. All regression models controlled for: Gender, School, and Age<br><sup>a</sup> Log dependent variable requires the transformation $100 * (2.7182^{\beta} - 1)$ [43] to interpret parameter estimate.<br><sup>b</sup> Elementary school category dropped by software due to incomplete information in dependent variables |  |  |  |  |

In the EMA model (i.e., prospective PA: model 1), youth whose ethnicity was categorized as visible minorities reported less PA (β = -0.911, 95% confidence interval [C.I.] = -1.677, -0.146, p–value = 0.024) in comparison with youth whose ethnicity was Canadian. This association was not found to be statistically significant in the retrospective PA model (i.e., retrospective PA: model 2). Similarly, youth who reported at least one parent having a university degree accumulated more EMA PA (β = 0.978, 95% [C.I.] = 0.308, 1.649, p–value = 0.006) in comparison with youth who reported that their parents had at least secondary school education. This association was not found to be statistically significant in the retrospective PA model. Following the same pattern, youth who engaged in at least one day of strength training reported more PA via EMAs (β = 0.932, 95% [C.I.] = 0.108, 1.755, p-value = 0.031) in comparison with youth who reported zero days of strength training. This association was not found to be statistically significant in the retrospective PA model.

In the retrospective PA model, youth who reported having at least one friend who is physically active were significantly associated with more PA (β = 0.741, 95% [C.I.] = 0.026, 1.458, p-value = 0.048) in comparison to youth who reported having zero physically active friends. This association was not found to be statistically significant in the EMA model.

## Discussion

This study was conducted by engaging youth citizen scientists using their own smartphones by utilizing a methodology that integrates ethical population health surveillance, integrated knowledge translation, and real-time behavioural interventions [32]. The primary purpose of this study was to ascertain if there was a significant difference between the duration of PA reported retrospectively using a modified version of three validated PA questionnaires and duration of PA reported prospectively using EMAs within the same cohort of youth aged 13-21 years. In addition, the study also assessed sociodemographic and contextual factors that are associated with duration of PA reported retrospectively vs. prospectively.

The primary finding was that there was a significant difference in PA reported retrospectively using the modified retrospective PA survey in comparison with PA reported prospectively using EMAs, with youth reporting more PA using prospective EMAs. In a similar study, although carried out on a cohort of adult participants [26], more PA was reported using a validated retrospective survey in comparison to EMAs. This discrepancy between adult and youth reporting of duration of PA needs further exploration. Evidence indicates that there are differences in how adults and youth accumulate PA [44], with youth engaging more in frequent habitual activities [45], resulting in intermittent PA [45], that could be better suited to capture via prospective EMAs. There is also some evidence that overestimation of PA using self-reported retrospective measures is more likely to occur in older adults [36]. More importantly, as this study was implemented via ubiquitous digital tools (i.e., smartphones), it is important to consider the level of digital literacy of youth, which is known to be higher than adults [46,47], could have played a role in youth having proficiency in reporting PA using digital EMAs.

According to the United Nations Educational, Scientific and Cultural Organization, “digital literacy is the ability to access, manage, understand, integrate, communicate, evaluate, and create information safely and appropriately through digital devices and networked technologies for participation in economic and social life. It includes competences that are variously referred to as computer literacy, ICT literacy, information literacy, and media literacy” [48]. In employing EMAs in prospective capture of duration of PA, it is important to consider the digital literacy of the participants to ensure that the methods of EMA deployment align with the level of digital literacy. In our study, we used citizen science approaches to engage with youth before EMA deployment to ensure that they can report PA with ease [26]. This engagement included relationship building with the schools and organizing presentation sessions, where youth had the opportunity to ask questions and even suggest potential changes to EMA deployment. For instance, although before deployment EMAs were set up to expire within an hour, after feedback from youth citizen scientists during the engagement sessions, we increased the time of expiry to maximize daily PA reporting opportunities [27]

As for the sociodemographic and contextual factors associated with duration of PA, this study indicates that there are several differences in associations between duration of PA reported retrospectively vs. prospectively. The retrospective PA model (Table 3: Model 2) depicted one significant association, where youth who reported having at least one physically active friend also reported more minutes of PA. This finding is consistent with previous quantitative and qualitative PA studies [49–51], where peer support was found to increase PA of youth. The EMA model (Table 3: Model 1) depicted three significant associations. Youth who reported at least one parent having a university degree in comparison with youth whose parents have high school or lower education, and youth who engaged in at least one day of strength training in comparison to youth who reported zero days of strength training reported more minutes of PA. Although not many studies have been carried out to ascertain factors that are associated with PA duration reported using EMAs by youth, these findings are consistent with existing evidence [52–55]. Finally, the EMA model also showed that visible minority youth reported lower duration of PA in comparison with youth who identified themselves as Canadian, a finding that is consistent with previous studies that have examined differences of PA among ethnicities [56,57].

More importantly, although our findings are consistent with existing evidence, the key finding here is the difference in sociodemographic and contextual factors that were associated with duration of PA reported retrospectively vs. prospectively. If we are to develop appropriate interventions to address global physical inactivity [58–60], it is critical to accurately understand the factors that determine PA accumulation. A clear difference between factors that are associated with PA reported retrospectively vs. prospectively by the same cohort of individuals shows that further investigation is needed to understand physical activity measurement, particularly in the digital age, where ubiquitous tools are available to obtain data in real-time [26,27,61].

There is considerable evidence that prospective EMAs are effective measures in estimating determinants and correlates of PA in real-time and real-world settings and their validity and reliability in measuring PA has been established [62,63]. EMAs reduce participant burden by using digital reminders/nudges that can be triggered on participants’ smartphones based on time, and location. EMAs can also be self-triggered by participants, which provides them the capacity to provide information that is tailored to their needs and circumstances [64,65]. EMAs are also known to reduce recall bias since participants do not need to recall their behaviours [17], a significant factor in improving PA measurement in real-world settings.

It is important to note that EMAs can transform how data are collected in the digital age, because they can be completed at participants’ convenience, and in collaboration between the researchers and the participants i.e., digital citizen science [26,66]. It is also important to consider the age cohort involved in data collection i.e., EMAs are more appealing to young participants with greater digital literacy [46,63].

Evidence also indicates that EMAs provide ecological validity of whether associations are significant in relation to typical settings of everyday life [67,68]. PA measurement using EMAs provide context, improves data validity through reduction of recall bias and data entry errors as participants are not required to retrospectively recall their behaviours [69,70]. EMAs reduce participant burden by using digital reminders/ nudges that can be triggered on participants’ phones based on predefined time and location [69,71]. In addition, EMAs can also be self-triggered by participants, which allows them some level of personalization to their needs [69,72]. One clear indication is that in the digital age, where smartphone usage is almost universal [73,74], it is critical to further explore usage of digital EMAs to capture PA across populations. This exploration is especially important due to PA’s role in minimizing non-communicable diseases [1,75]. As global PA patterns are consistently reported to educate the public, and to inform policies to prevent non-communicable diseases [76–81], and as it is important to accurately capture PA patterns, digital citizen science could play an important role in ethical surveillance of PA [27].

## Strengths and Limitations

Strengths of this study include further enhancing our understanding regarding digital tools and methodologies in reporting health behaviours. It will contribute to new evidence given the digitization of the surveys themselves, but also because they capture prospective data at participants’ convenience. Such data are collected by researchers in real-time, who can adjust surveys if needed or send prompts in real-time. In addition, the combination of three validated surveys to come up with a modified retrospective survey suited for smartphone-based deployment adds to the strength of this study in this digital age where smartphones have become ubiquitous. Limitations of this study include subjective surveys within cross-sectional design, and future research should combine EMA measures with objective data collection using longitudinal designs.

## Conclusion

Physical inactivity is the fourth leading cause of mortality globally, and it is critical to understand patterns of PA using rigorous and validated tools. The findings of this study show the importance of using prospective EMAs to capture PA, which is particularly relevant in the digital age, where ubiquitous devices provide us with real-time engagement capabilities. More importantly, digital citizen science can transform how we measure behaviours using citizen-owned ubiquitous digital tools to support prevention and treatment of non-communicable diseases.

## Data Availability

Data are available from the University of Regina’s Institutional Data Access / Ethics Committee for researchers who meet the criteria for access to confidential data.

## Acknowledgments

The authors acknowledge the entire Digital Epidemiology and Population Health Laboratory team (DEPtH) for their unwavering support. Authors also acknowledge the Canadian Institute of Health Research for their support to the DEPtH Lab and the Smart Platform.

